# Soap versus sanitiser for preventing the transmission of acute respiratory infections: a systematic review with meta-analysis and dose-response analysis

**DOI:** 10.1101/2020.07.22.20160432

**Authors:** Tammy Hoffmann, Mina Bakhit, Natalia Krzyzaniak, Chris Del Mar, Anna Scott, Paul Glasziou

## Abstract

**Objective:** To compare the effectiveness of hand hygiene using alcohol-based hand sanitiser to soap and water for preventing the transmission of acute respiratory infections (ARIs), and assess the relationship between the dose of hand hygiene and the number of ARI, influenza-like illness (ILI), or influenza events.

**Methods:** Systematic review of randomised trials that compared a community-based hand hygiene intervention (soap and water, or sanitiser) with a control, or trials that compared sanitiser with soap and water, and measured outcomes of ARI, ILI, or laboratory-confirmed influenza or related consequences. Searches were conducted in CENTRAL, PubMed, Embase, CINAHL and trial registries (April 2020) and data extraction completed by independent pairs of reviewers.

**Results:** Eighteen trials were included. When meta-analysed, three trials of soap and water versus control found a non-significant increase in ARI events (Risk Ratio (RR) 1.23, 95%CI 0.78-1.93); six trials of sanitiser versus control found a significant reduction in ARI events (RR 0.80, 95%CI 0.71-0.89). When hand hygiene dose was plotted against ARI relative risk, no clear dose-response relationship was observable. Four trials were head-to-head comparisons of sanitiser and soap and water but too heterogeneous to pool: two found a significantly greater reduction in the sanitiser group compared to the soap group; two found no significant difference between the intervention arms.

**Conclusion:** Adequately performed hand hygiene, with either soap or sanitiser, reduces the risk of ARI virus transmission, however direct and indirect evidence suggest sanitiser might be more effective in practice.

## 1. Introduction

Acute respiratory infections (ARI) cause a substantial annual health burden, and much more so in the current COVID-19 pandemic. Hand hygiene is one effective and low-cost intervention which reduces the transmission of ARIs [1] and is applicable in all countries and all settings. However, important questions for policy and practice are the “dose-response” of hand hygiene, and relative effectiveness of different materials (alcohol-based hand sanitiser; soap and water). This systematic review aimed to address these questions.

## 2. Methods

### 2.1. Inclusion criteria and study source

Our recent systematic review and meta-analysis of physical interventions to interrupt or reduce the spread of respiratory viruses [2] (an update of the 2011 review [3]) aimed to synthesise all randomised controlled trials of several physical interventions (including hand hygiene) which measured outcomes of ARI, influenza-like illness (ILI), or laboratory-confirmed influenza (influenza) or related consequences (e.g. absenteeism). For the current systematic review, trials were eligible if they compared a hand hygiene intervention with a control, or compared hand sanitiser with soap and water. Trials in healthcare settings were excluded. We also screened a new Cochrane review of rinse-free handwashing in school and pre-school children for possible eligible studies [4].

### 2.2. Data extraction

Data were extracted by two authors (MB, NK) independently on: volume or weight of material (e.g. sanitiser or soap) used per person per day, and number of handwashes per person per day. When not reported directly, we estimated usage where possible (see Table 1). For estimation purposes, we used data on the average amount of material used per person per handwash as reported; if data were not reported, we assumed 0.035 grams of soap or 1.5ml of liquid used per handwash [5]. The following data were extracted from the parent systematic review [2]: 1) study characteristics; 2) risk of bias assessments; 3) type of handwashing intervention(s) (e.g. soap, sanitiser, gel); and 4) risk ratios (RR), log RR, and standard error values for ARI or ILI or influenza (including the outcome with most events from each study).

**Table 1.**
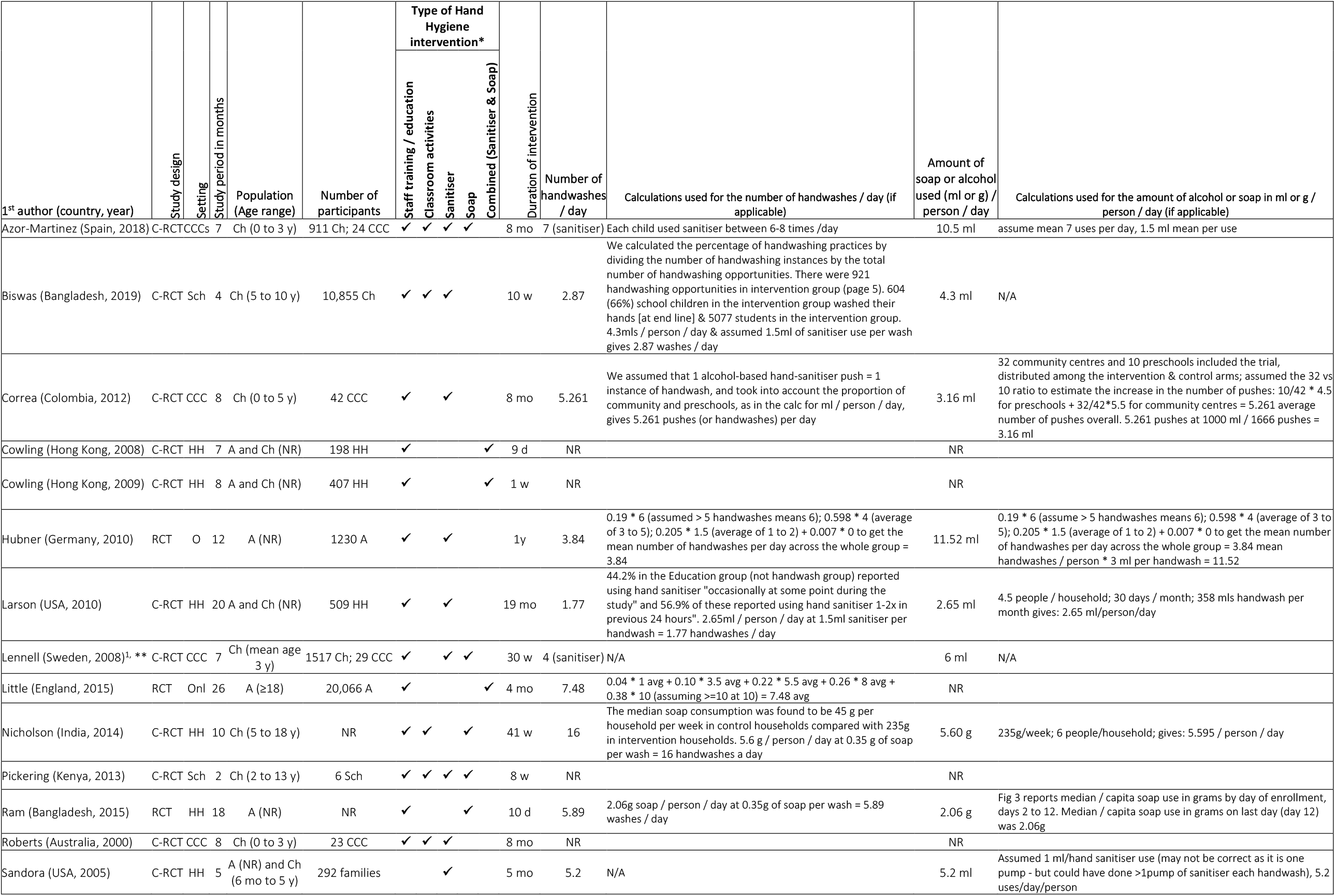
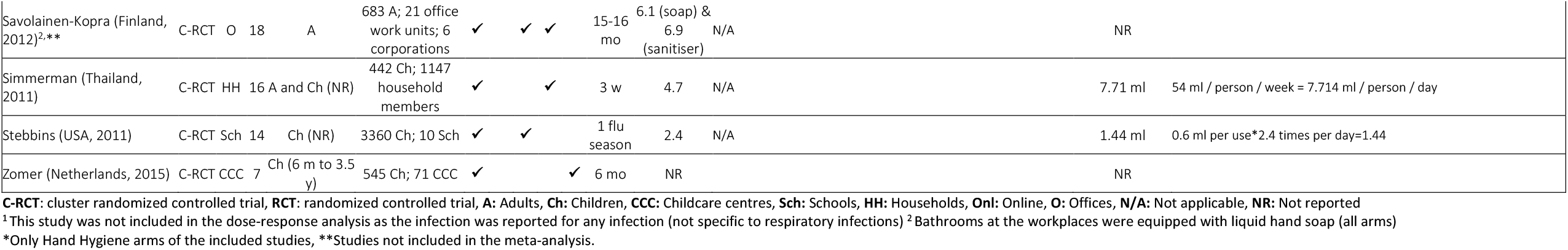
Characteristics of included studies.

### 2.3. Data analysis

To assess the relationship between handwashes per person per day and the number of ARI or ILI or influenza events, we conducted the following analyses: 1) only studies whose number of handwashes could be estimated (regardless of the type of handwash material), subgrouped by the type of handwash material (soap vs sanitiser vs combination of sanitiser and soap) and 2) all studies (whether or not the number of handwashes could be estimated), subgrouped by the type of handwash material (soap vs sanitiser vs combination of sanitiser and soap). We used a Chi^2^ test to test for subgroup interactions. Meta-analyses were conducted using *Review Manager 5*.

## 3. Results

The PRISMA flowchart (Fig 1) shows the number of trials identified from the updated 2020 systematic review [2], the original 2011 review [3], and additional sources. Eighteen trials were assessed as eligible; four were head-to-head comparisons of hand sanitiser and soap and water [6-9] and 16 compared hand hygiene with a control [6, 8, 10-23]. Table 1 presents study characteristics.

**Figure 1.**
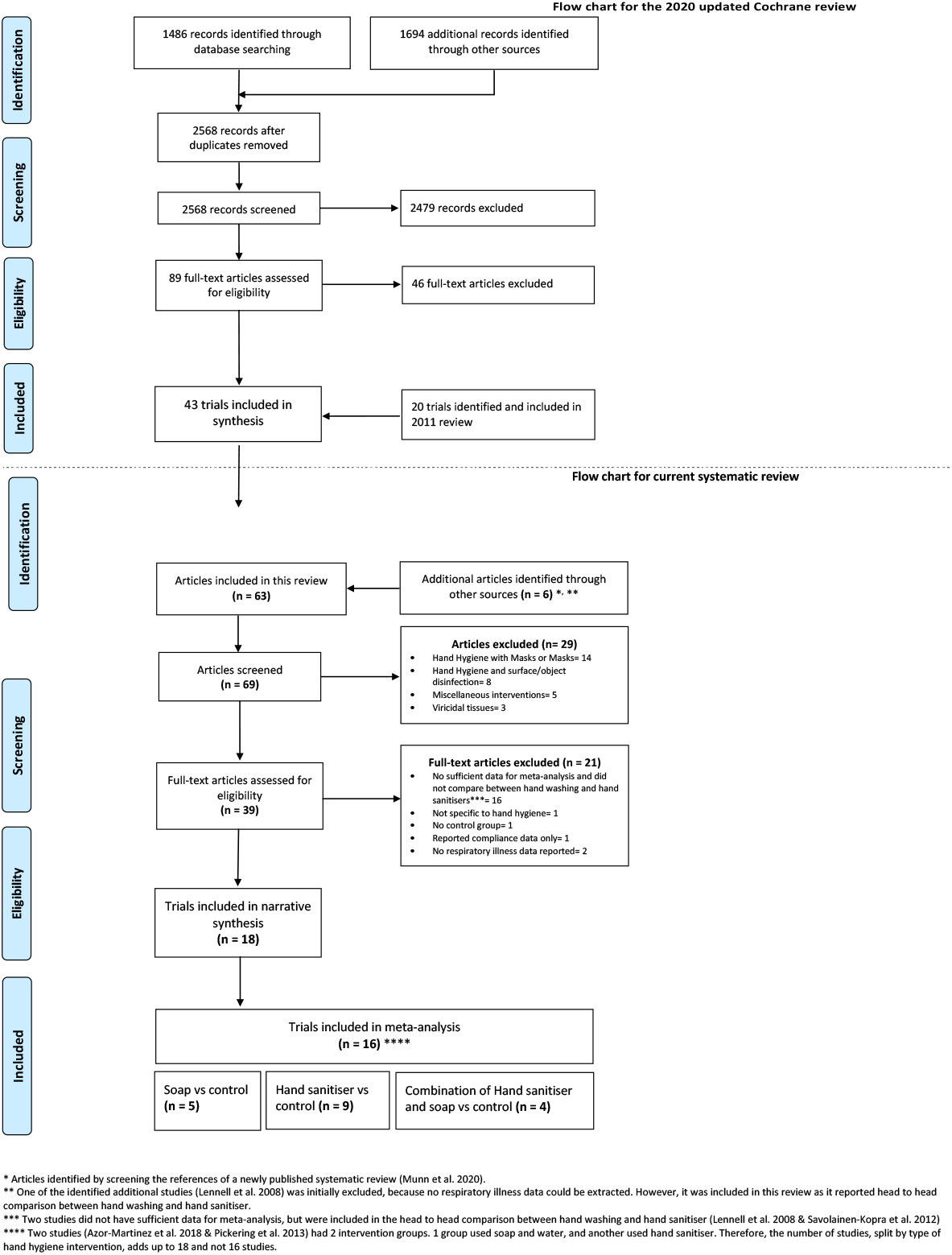
PRISMA Flow Chart.

### 3.1. Trials of hand sanitiser or soap and water versus control

Combining the three trials of soap and water hand hygiene versus control found a non-significant increase in ARI events: risk ratio 1.23 (95% CI 0.78 to 1.93) but with high heterogeneity (Figure 2; Appendix A.1 shows forest plot for all trials, regardless of whether number of handwashes could be estimated). Combining the six trials of hand sanitiser versus control found a significant reduction in ARI events: risk ratio 0.80 (95% CI 0.71 to 0.89), providing some indirect evidence in favour of hand sanitiser.

**Figure 2.**
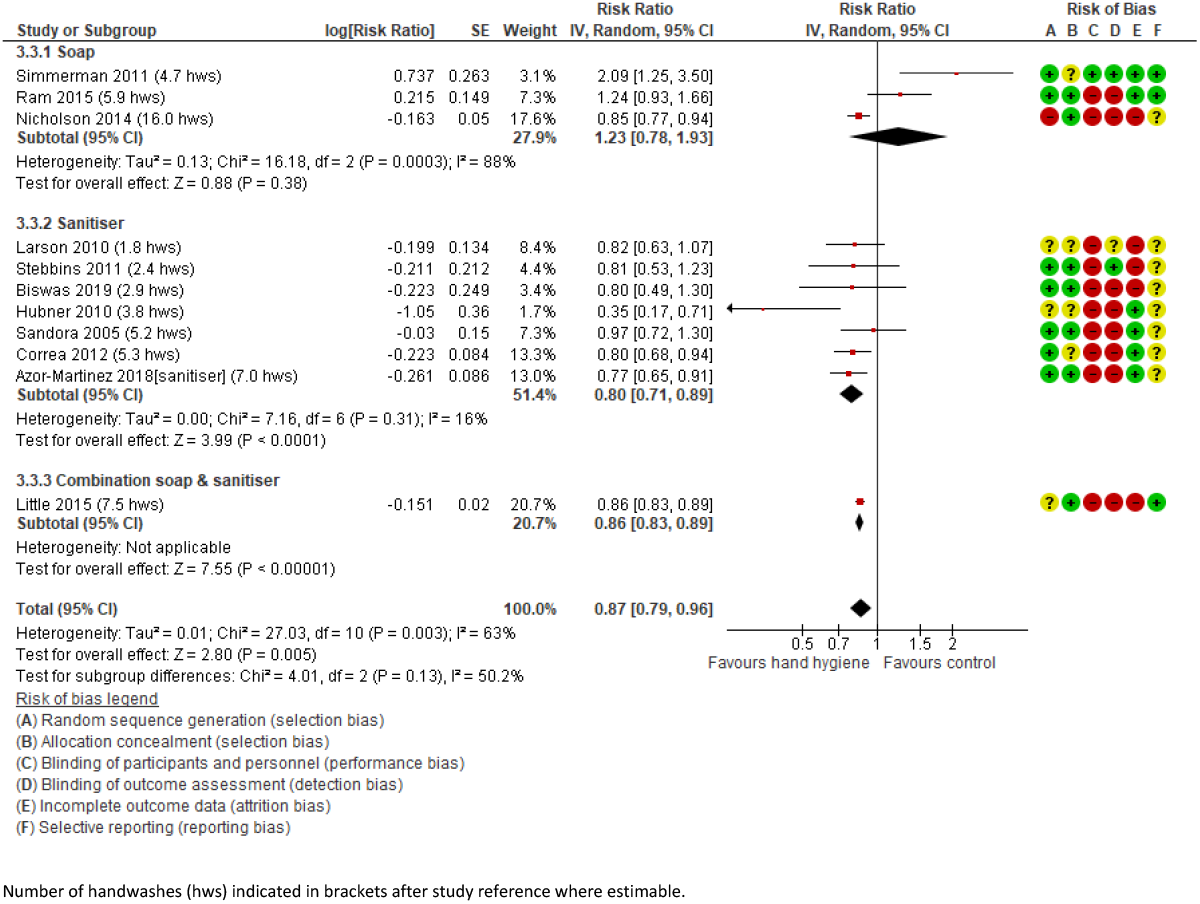
**Forest plot of meta-analysis of studies whose number of handwashes could be estimated, subgrouped by the type of handwash material (soap vs sanitiser vs combination of sanitiser and soap)**

### 3.2. Dose-response relationship: hand hygiene frequency versus risk of respiratory infection (ARI, ILI or influenza)

Eleven of the trials provided sufficient information to estimate the dose of hand hygiene, which we converted to number of hand hygiene events per day. Plotted against the relative risk of ARIs, there is little dose response relationship evident for hand sanitiser (Figure 3). There are only three studies solely of soap and water hand hygiene, making a dose-response analysis impossible. The difference in effectiveness between hand sanitiser and soap and water does not appear to be explained by a difference in frequency. The cluster randomised trial by Little and colleagues [16] primarily used soap and water but also offered participants free hand sanitiser; only 18% report collecting the sanitiser.

**Figure 3.**
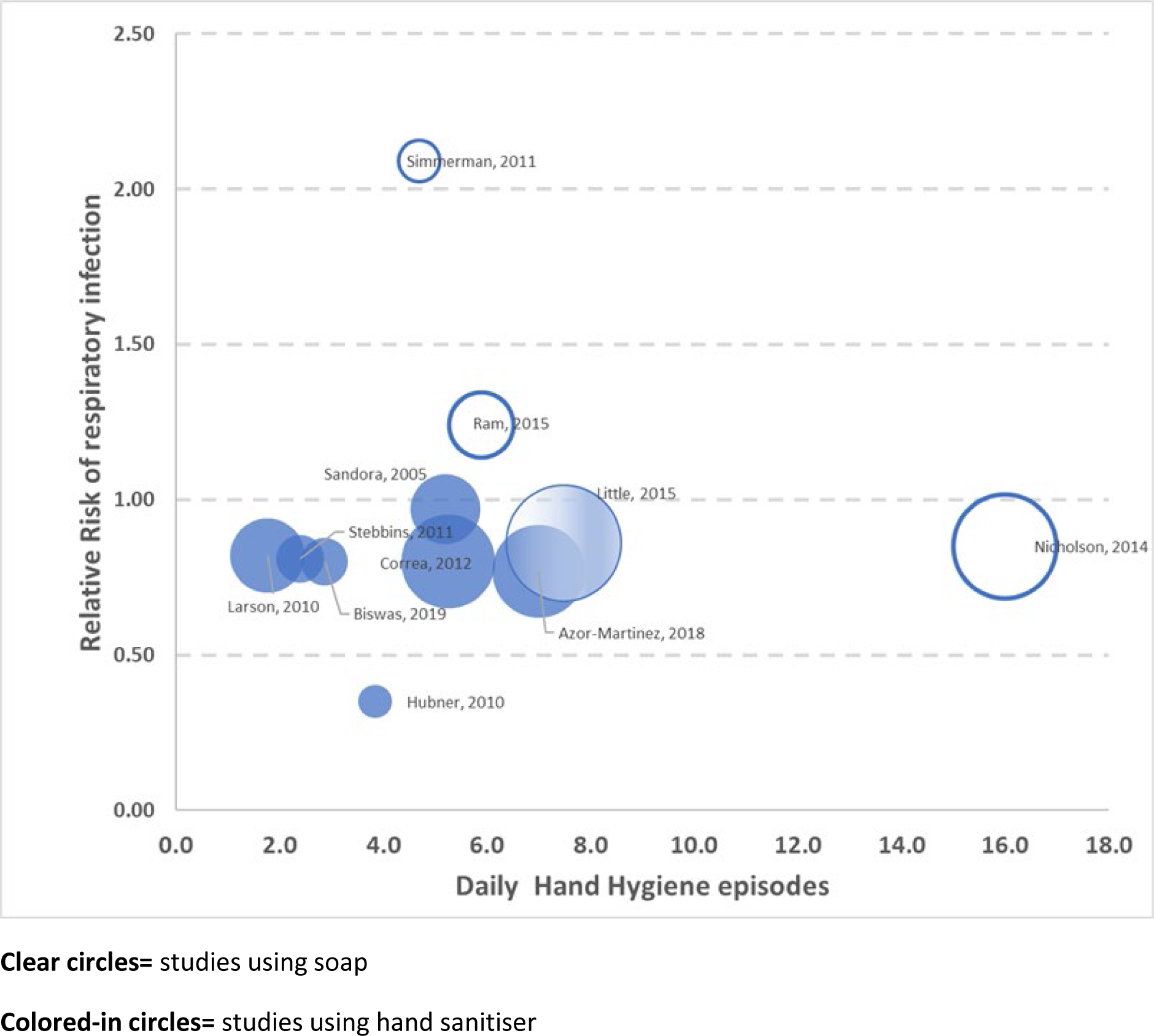
Hand hygiene frequency (‘dose’) versus risk of respiratory infection (ARI, ILI or influenza)

### 3.3. Head-to-head trials of hand hygiene with hand sanitiser vs with soap and water

Four trials directly compared hand sanitiser with soap and water: two in childcare centres, one at a primary school, and one in workplaces. In a cluster randomised trial of children and staff in Swedish childcare centres, those at centres who were randomised to use an alcohol-based oily disinfectant gel (70% ethanol) after regular hand washing had a reduction in absenteeism rate of 12% (95% CI 4% to 20%) compared to control centres which used only soap and water [7]. The 3-arm cluster randomised trial of 24 childcare centres in Spain - educational and hand hygiene measures (one with soap and water; another with hand sanitiser) and a control group found children in the sanitiser group had a 13% lower (95% CI 6% to 28%) risk of respiratory infection than children in the soap and water group [6].

In Kenya, a cluster randomised trial assigned two primary schools to receive a handwashing with soap and water intervention, two to receive a sanitiser intervention, and two were a control [8]. Compared to control group students, both intervention groups had a reduction in observed rhinorrhea (RR 0.77, 95% CI 0.62-0.95 for both sanitiser vs control and soap vs control). No significant differences between the sanitiser and soap groups were observed for respiratory outcomes. The 3-arm trial in six companies in Finland randomised workplaces to equip workplace bathrooms with liquid hand soap (soap and control arms) or alcohol-based hand rub [9]. Participants in the intervention arms also received guidance on additional strategies for limiting infection transmission. Before the onset of the 2009 influenza pandemic (and the subsequent national hand hygiene campaign), a statistically significant (p = 0.002) difference in the infection episodes was observed between the control (6.0 per year) and the soap-and-water arm (5.0 per year), but not between the control and the alcohol-rub arm (5.6 per year). Neither intervention had an effect on work absenteeism.

## 4. Discussion

Based on both indirect and direct (head-to-head) trials, hand hygiene using alcohol-based hand sanitiser appears more effective at reducing ARI transmission than hand hygiene using with soap and water, with the difference in effect not explained by the difference in frequency of hand hygiene. The apparent greater effectiveness of hand sanitiser may be explained by its greater convenience, less time required to perform hand hygiene, more sustained compliance with hand hygiene, and less irritation to the skin [24].

Limitations of this review are that conclusions are mostly from indirectness evidence, with direct evidence available from only four head-to-head trials, and that it was not possible to estimate the dose of hand hygiene for some trials.

A recent Cochrane review of the effect of rinse-free handwashing, compared to traditional hand hygiene, on absenteeism for ARI in preschool and school children reported a significant reduction in absenteeism of 9 days per 1000 available days for children in the rinse-free group, with the results coming from six randomised trials [4]. The effectiveness of handwashing with materials other than sanitiser or soap and water, such as ash, which may be used in low-income countries has mostly been examined in observational studies with uncertain effects [25].

## 5. Conclusions

Hand hygiene has a modest but important role in reducing the transmission of viral respiratory infections. Adequately performed hand hygiene, with either soap or sanitiser, reduces the risk of acute respiratory virus transmission. However, from both the direct and indirect comparisons in this review, sanitiser appears more effective in practice. While further head-to-head randomised trials are warranted, the current evidence appears sufficient to promote the use of hand sanitiser as the primary means for many everyday situations.

## Data Availability

No additional data available

## Abbreviations

(ARI): Acute respiratory infections
(ILI): Influenza-like illness
(RR): Risk Ratio
(CI): Confidence interval

## Contributors

TH, PG, and CDM conceived the study. MB and NK screened studies for inclusion in this review and extracted the data. TH, AMS, PG and CDM analysed the data. TH wrote the first draft. All authors approved and revised the final manuscript.

## Competing interests

None declared.

## Funding

This research did not receive any specific grant from funding agencies in the public, commercial, or not-for-profit sectors.

## Ethical approval

Ethical approval was not required.

## Acknowledgements

We would like to acknowledge the authors of the 2020 Cochrane review update (L Al-Ansary, G Bawazeer, E Beller, J Clark, J Conly, E Dooley, E Ferroni, T Jefferson, S Thorning, M van Driel, M Jones).

## Appendix A. Supplementary data

Appendix A.1 – Forest plot of meta-analysis of all studies (regardless of whether the number of handwashes could be estimated) regardless of the type of handwash material (soap vs sanitiser vs combination of sanitiser and soap)

## Notes

### Competing Interest Statement

The authors have declared no competing interest.

### Funding Statement

No funding

### Author Declarations

No Ethics approval required

